# One in seven pathogenic variants can be challenging to detect by NGS: An analysis of 450,000 patients with implications for clinical sensitivity and genetic test implementation

**DOI:** 10.1101/2020.07.22.20159434

**Authors:** Stephen E. Lincoln, Tina Hambuch, Justin M. Zook, Sara L. Bristow, Kathryn Hatchell, Rebecca Truty, Michael Kennemer, Brian H. Shirts, Andrew Fellowes, Shimul Chowdhury, Eric W. Klee, Shazia Mahamdallie, Megan H. Cleveland, Peter M. Vallone, Yan Ding, Sheila Seal, Wasanthi DeSilva, Farol L. Tomson, Catherine Huang, Russell K. Garlick, Nazneen Rahman, Marc Salit, Stephen F. Kingsmore, Matthew J. Ferber, Swaroop Aradhya, Robert L. Nussbaum

## Abstract

**Purpose:** To evaluate the impact of technically challenging variants on the implementation, validation, and diagnostic yield of commonly used clinical genetic tests. Such variants include large indels, small CNVs, complex alterations, and variants in low-complexity or segmentally duplicated regions.

**Methods:** An interlaboratory pilot study used novel synthetic specimens to assess detection of challenging variant types by various NGS-based workflows. One well-performing workflow was further validated and used in clinician-ordered testing of more than 450,000 patients.

**Results:** In the interlaboratory study, only two of 13 challenging variants were detected by all 10 workflows, and just three workflows detected all 13. Limitations were also observed among 11 less-challenging indels. In clinical testing, 21.6% of patients carried one or more pathogenic variants, of which 13.8% (17,561) were classified as technically challenging. These variants were of diverse types, affecting 556 of 1,217 genes across hereditary cancer, cardiovascular, neurological, pediatric, reproductive carrier screening, and other indicated tests.

**Conclusion:** The analytic and clinical sensitivity of NGS workflows can vary considerably, particularly for prevalent, technically challenging variants. This can have important implications for the design and validation of tests (by laboratories) and the selection of tests (by clinicians) for a wide range of clinical indications.

## INTRODUCTION

Clinical genetic tests based on next-generation sequencing (NGS) are increasingly used to aid diagnosis and inform patient care.^1^ Specific guidelines are available for the clinical application of this technology,^2–4^ and many NGS-based tests that detect single-nucleotide variants (SNVs) and small insertions and deletions (indels) in relatively accessible parts of patients’ genomes have been implemented. NGS has also been extended to detect copy-number variants (CNVs, also called del/dup events)^5–8^ although these methods are less uniformly implemented.

However, conventional short-read NGS methods have well-known limitations that can allow certain technically challenging variants — such as large indels, small CNVs, and complex alterations — to remain undetected, at least without the application of specialized bioinformatic and biochemical methodologies.^9–11^ Even simple SNVs and indels in segmentally duplicated (segdup) or low-complexity genomic regions can present substantial challenges.^12–15^

The impact of such technically challenging variants on the implementation, validation, and diagnostic yield of commonly used clinical genetic tests has not yet been thoroughly described. In this study, we examined the spectrum of pathogenic germline variants uncovered during germline genetic testing of specific genes or multi-gene panels for hereditary cancer, cardiovascular, neurological, and pediatric disorders, reproductive carrier screening, and other clinical indications. To accomplish this, we first conducted an interlaboratory pilot study, evaluating 10 different NGS laboratory workflows using novel, synthetic, positive controls containing variant types known to be challenging for NGS. One of these workflows was implemented and scaled-up in a high-volume clinical testing laboratory, and the sensitivity of this workflow was further evaluated using additional synthetic, reference, and clinical specimens. Finally, we examined the attributes of pathogenic variants reported using this workflow in daily practice.

## METHODS

### Pilot study

Synthetic plasmids containing variants of interest were constructed as described previously^16^ by SeraCare (Gaithersburg, MD). These novel plasmid designs included pathogenic variants, previously observed in patients, presenting specific technical challenges for NGS (Figure 1, Table S1). The plasmids were titrated into genomic DNA from the Genome in a Bottle (GIAB) GM24385 (HG002) cell-line^17^ at appropriate concentrations for the synthetic variants to appear heterozygous. These samples were provided to collaborating laboratories who sequenced them using various NGS workflows (Table 1, Table S2), each of which used a different bioinformatics pipeline. Four hybridization-based targeting chemistries, whole genome sequencing, and amplicon sequencing were represented. Most workflows used Illumina (San Diego, CA) sequencing and one used Ion Torrent (Thermo Fisher, Waltham, MA). Read-level data were examined using the Integrative Genomics Viewer (IGV).^18^ The construction methodology used for these controls created artifactual split-read and copy-number signals at known locations that were ignored (Table S3). Additional details are available online.^19^

**Table 1.**
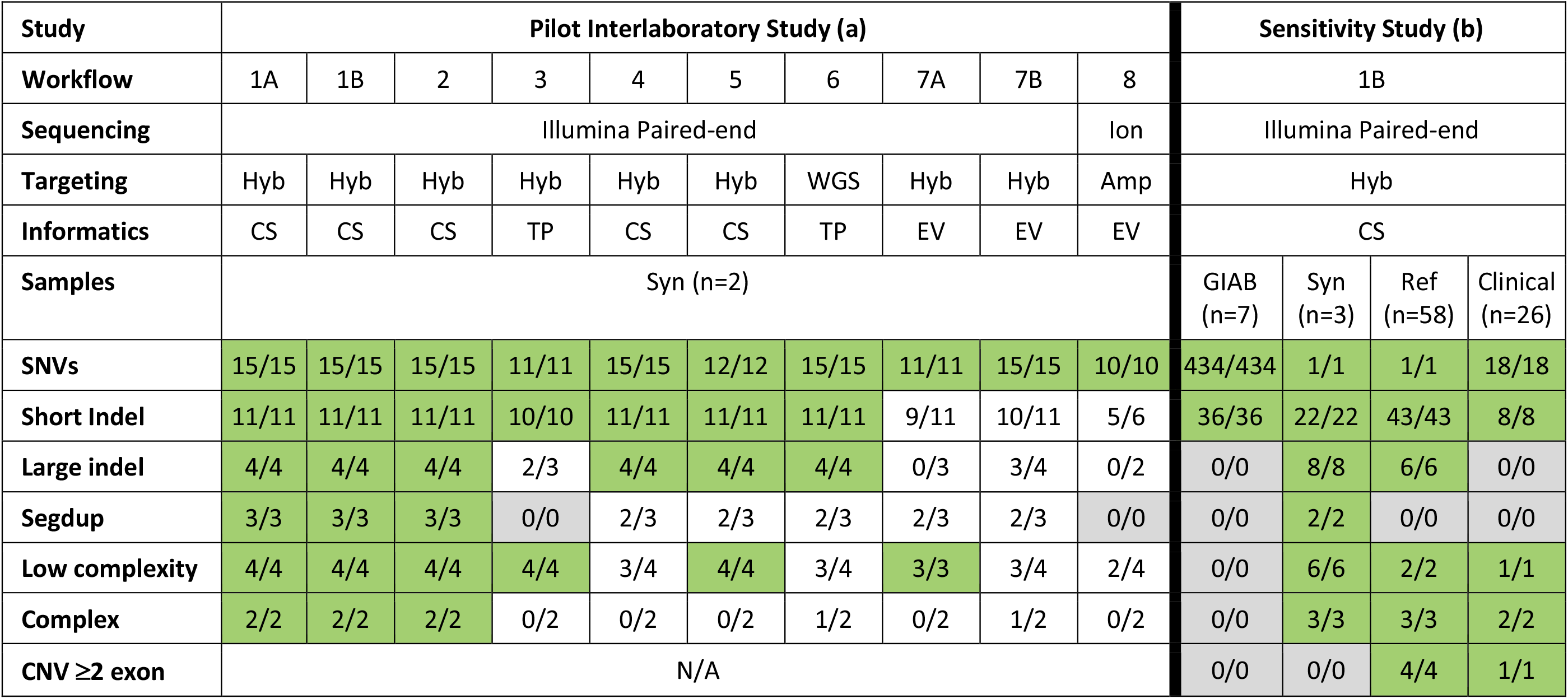
Proof of concept and sensitivity study results. For the interlaboratory pilot study (a), performance of ten NGS workflows is shown on variants in the synthetic controls. In each data cell, the denominator is the number of variants within each assay’s target regions and the numerator is the number of these variants that were detected. Green cells indicate 100% sensitivity. White cells indicate an observed limitation. Gray cells indicate that no study variants were present in regions interrogated by the assay. Details of each of the 10 workflows and the variants are provided in Table S2. Workflow 1A had previously detected these variants in patients, and was included to validate the synthetic controls. For the sensitivity study (b), performance is shown for variants in samples from each source. Positive controls with technically challenging variants were difficult to obtain, requiring a large number of reference specimens and additional synthetic controls. A list of samples used in this study is provided in Table S5. **Abbreviations**: Amp, amplicon sequencing; CNV; copy number variant; CS, custom software; EV, software provided by the sequencing equipment vendor; GIAB, Genome in a Bottle; Hyb, hybridization capture; Indel; insertion or deletion; Ion, Ion Torrent; N/A, not applicable; NGS, next-generation sequencing; Ref, reference specimens from public biobanks; Segdup, segmental duplication; SNVs, single nucleotide variants; Syn, synthetic controls; TP, third-party software; WGS, whole genome sequencing.

**Figure 1.**
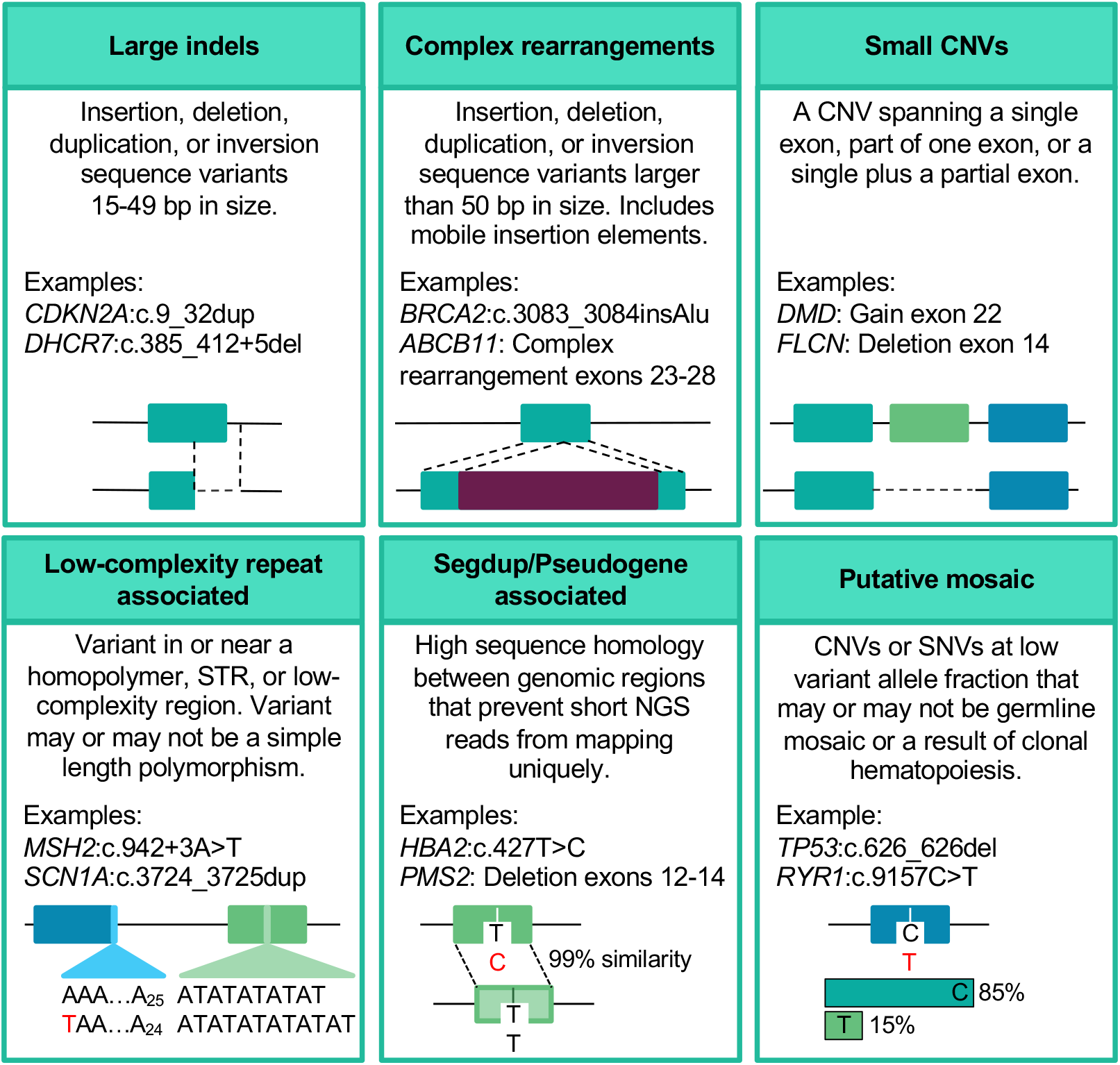
Technically challenging variant types. Variants were categorized as being technically challenging, or not, based on these six criteria. Note that some variants could be considered challenging for multiple reasons (e.g., a single-exon deletion within a segmentally duplicated region). Examples provided are variants observed in the prevalence analysis. **Abbreviations**: CNV, copy number variant; Indels, insertions or deletions; NGS; next-generation sequencing; Segdup, segmental duplication; SNVs, single nucleotide variants; STR; short tandem repeat.

### NGS methods for detecting a broad spectrum of genetic variation

In our sensitivity and prevalence studies, NGS was performed as described previously^7^ with improvements (Figure 2). In brief, targeted-capture libraries were created using customized probes from Integrated DNA Technologies (Coralville, IA), Roche Sequencing Solutions (Pleasanton, CA), or Twist Bioscience (South San Francisco, CA). Extra probes were included for sites that otherwise had low coverage. Paired-end sequencing (2×150) was performed on the Illumina NovaSeq 6000 to at least 300x average depth per sample (typically much higher) and at least 50x at each targeted position. In rare cases, specific sites with 20-49x coverage could also be accepted following additional data review and lab director approval.

**Figure 2.**
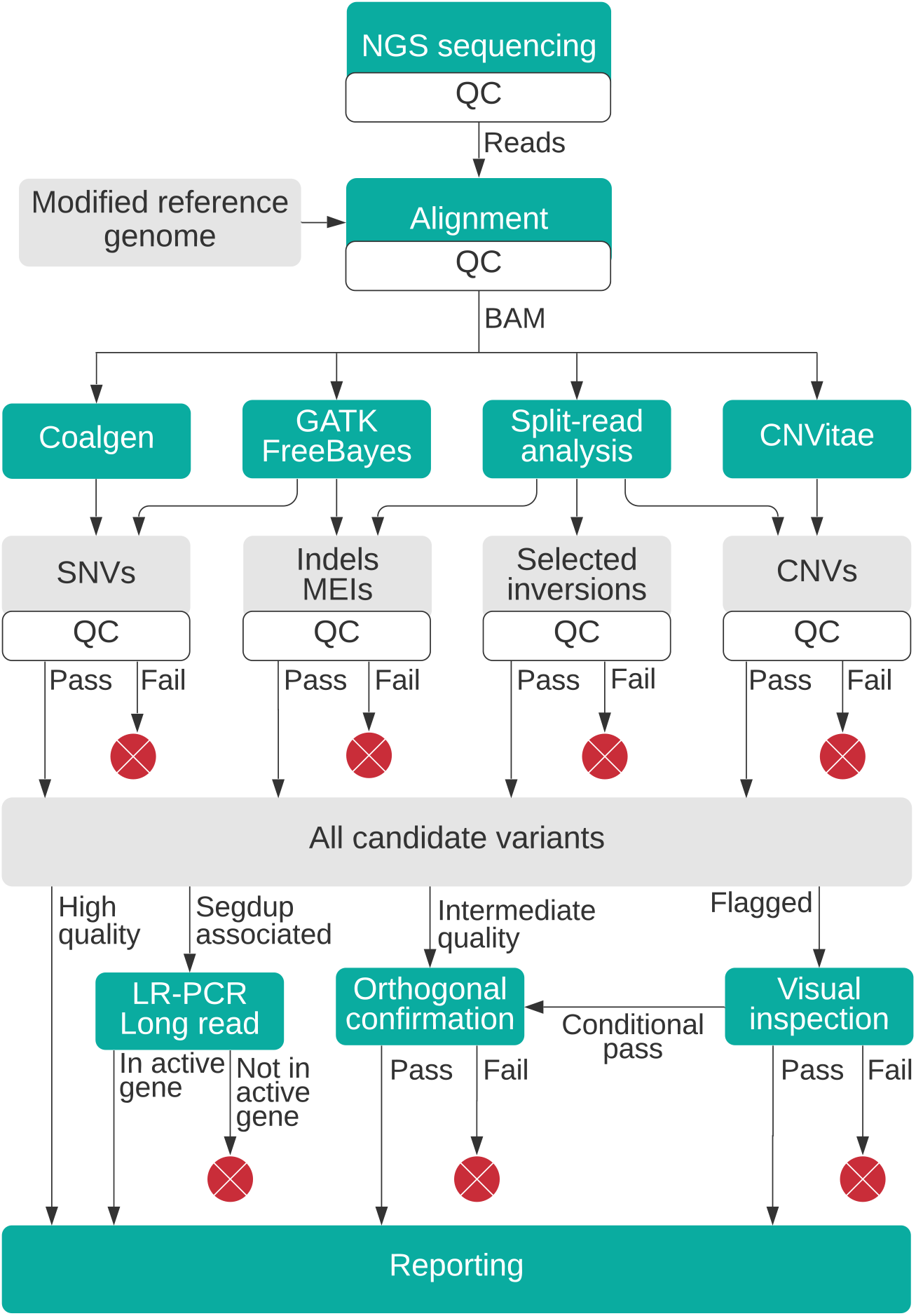
Variant calling process used in prevalence study. An NGS workflow designed to detect a wide variety of variant types was used in our prevalence study. NGS reads are aligned to a modified reference genome and multiple variant callers are then applied. Follow-up assays are used both to confirm potential false positives, to determine the exact sequence of complex variants, and to resolve the location of variants within segmental duplications. For details see Methods. **Abbreviations**: CNVs, copy number variants; GATK, Genome Analysis Toolkit; Indels, insertions or deletions; LR-PCR, long-range polymerase chain reaction; MEIs, mobile element insertions; NGS, next-generation sequencing; QC, quality control; Segdup, segmental duplication; SNVs, single nucleotide variants.

Reads were aligned with NovoAlign (Novocraft Technologies, Selangor, Malaysia) to the GRCh37 reference genome^20^ modified to improve variant detection in segdups. For regions where a duplicated copy exists that was both (a) not in GRCh37 and (b) sufficiently diverged from the main locus to allow 2×150 reads to map uniquely, we added the paralogous sequence to the reference genome. This reduces reads from the paralog mapping to the main locus (and vice versa). An example includes *PRSS1*, which has a polymorphic pseudogene not fully represented in GRCh37. Alternatively, for genes where the paralog is both (a) present in GRCh37 and (b) nearly identical to the main locus (i.e., situations where mismapping cannot be prevented), the paralogous regions were replaced with N’s in the reference genome to force NGS reads to map to a single location. In these cases, heterozygous variants from either locus would appear at an average allele fraction of 25%. Examples include *PMS2* (exons 12-15), *NEB* (exons 83-103), and *SMN1*/*SMN2*. When a potential pathogenic variant was suspected in these cases, a secondary assay using long-range PCR and long-read sequencing (Pacific Biosciences, Menlo Park, CA) was used as needed to determine in which locus a variant was present.

Sequence variants were called using a collection of algorithms, primarily the Genome Analysis Toolkit (GATK) Haplotype Caller.^21,22^ Freebayes^23^ was also used to improve analytic sensitivity for variants at low-allele fractions and Coalgen^7^ was used to detect specific homopolymer-associated variants. CNVs were called using a combination of read-depth analysis by CNVitae as well as split-read analysis.^7,8^ Split-read analysis also allowed the detection of large indels (including mobile element insertions) and copy-neutral structural variants with breakpoints in targeted regions; these variants can evade both read-depth and GATK-based detection.

Clinically significant variants at risk of being false positives were identified and confirmed using an orthogonal assay as described previously.^24^ A separate assay involving PCR and long-read sequencing was implemented for triplet repeat expansions in *FMR1*.^25^

### Sensitivity Study

Herein, we describe one sensitivity assessment of this workflow using a particular gene panel (Table S4) that exercises all of the workflow’s components. This study used GIAB samples^17^ with reference data version 3.3.2. Only variants that were both annotated as high-confidence calls by the GIAB Consortium and within our assay’s reportable target regions were included in this study. Benign polymorphisms meeting these criteria were abundant in the GIAB reference data and were included (clinical interpretation was not considered). Additional reference samples containing specific variants of interest were obtained (Table S5), and clinical specimens were included for which test results from an independent laboratory (Myriad Genetics, Salt Lake City, UT) were available. Finally, we utilized the synthetic controls described above and additional synthetic variants (Table S6). Unlike the pilot study, these additional variants were not known to be previously detected. All samples were processed in a blinded manner.

### Prevalence analysis

A consecutive series of patients receiving gene panel testing by Invitae (San Francisco, CA) between June 2018 and March 2020 was retrospectively analyzed. Exome-sequencing based tests were excluded owing to methodological differences with panel testing. The specific genes analyzed for each patient were a subset of those assayed, chosen by ordering clinicians. Variants were clinically classified using Sherloc, a framework based on the American College of Medical Genetics and Association for Molecular Pathology guidelines.^26,27^ Only pathogenic and likely pathogenic variants were included in our prevalence study – variants of uncertain significance and benign variants were excluded. Variants were categorized as technically challenging, or not, using specific criteria (Figure 1, Supplemental Methods).

### Ethics Statement

Our study protocol 20161796 was approved by the Western Institutional Review Board. Reference specimens were used under the terms of their respective material transfer agreements.

## RESULTS

### Interlaboratory pilot study

An interlaboratory study was conducted to both reinforce our understanding of the impact of different NGS methodologies on challenging variant types, and to evaluate whether synthetic positive controls are a useful tool for the development and validation of methods to detect such variants. In this study, all 10 NGS workflows at collaborating laboratories were able to sequence and analyze the synthetic control mixtures, demonstrating compatibility of the synthetic approach with various NGS biochemistries. However, only two of the 13 challenging variants (as defined in Figure 1) were detected by all 10 workflows, and just three workflows detected all 13 (Tables 1, S1, S2). Additionally, three of the 11 other indels (which were considered less challenging) were missed by some workflows.

Manual review using IGV demonstrated that evidence of the missed variants was visible in most of the raw data sets, indicating that the sensitivity limitations were largely bioinformatic in nature. IGV review of data from synthetic controls and patient specimens containing the same variants showed similar challenges including artifacts, misalignments, clipped reads, stutter, and deviations from 50:50 allele fractions (Figure S1). The amplicon sequencing workflow (number 8) was an exception however, as five of the 12 targeted indels were false negatives because (a) the variant altered a PCR primer binding site, (b) the variant was near an amplicon boundary, interfering with alignment, or (c) the variant caused a substantial increase in amplicon size, which the biochemistry could not accommodate. This sequencing platform also exhibited its characteristic limitation with both of the homopolymer-associated variants in this study.^19^Many, but not all, of the sensitivity limitations identified in this study were already known to the collaborating laboratories. Further review of the workflows’ components (Table S2) identified probable root causes of these sensitivity limitations and indicated that these limitations would likely apply to patient specimens and to other variants with similar properties (not just to the specific variants in this study)^19^. Our review also suggested workflow modifications that could be implemented to potentially improve performance. Overall, we determined that synthetic controls were an informative and valid tool for many challenging variant types.

### Sensitivity study

One NGS workflow (Figure 2) from the pilot study was used in our prevalence analysis (below). Its sensitivity was further evaluated using a methods-based approach^2–4,28^ rather than a gene- or variant-based approach, which was the only practical option considering the large number of genes and variants targeted. In such studies, positive control specimens containing a diversity of variants are obtained, and the ability to detect these variants is measured by class. This particular study utilized 94 specimens containing 601 independently characterized positive control variants in 47 genes of interest. All 601 were correctly detected, demonstrating 100% observed sensitivity (Table 1). We found that the manner in which the various specimen types contributed to this study varied considerably, with significant implications for the evaluation of methods to detect challenging variants which we elaborate on here.

The seven Genome in a Bottle (GIAB) samples, for example, contributed most (470/601, 78.2%) of the study variants, although these had limited clinical or methodological relevance. The vast majority (92%) were SNVs, while pathogenic variants in these 47 genes are often indels or CNVs.^7,29^ Moreover, none of the 36 GIAB indels were greater than 5 base-pairs (bp) in size, and many challenging genomic locations (e.g., the pseudogene-associated exons of *PMS2*) did not have any high-confidence calls in the GIAB 3.3.2 data. (Newer GIAB data sets may improve this particular limitation, however, as shown below). The GIAB samples do contain additional variant types^30^ (CNVs and structural variants) but these were not located in or near our targeted genes and were not useful for measuring sensitivity of this assay. Indeed, none of the 470 GIAB variants met our definition of technically challenging.

To increase the number of clinically important variant types, we included 58 additional reference samples (Table S5) and 26 clinical specimens. Unlike GIAB, each of these contributed only one or two independently characterized variants to our study. Nevertheless, this set added 60 indels, ten of which were larger than 5 bp, and nine CNVs. Most importantly, it provided 14 variants that met our definition of technically challenging (Figure 1).

However improved, these variant counts remained small, particularly considering the multiple, diverse challenging variant subtypes. We thus added synthetic controls to our study, which have the advantage of including multiple variants of interest in each DNA sample. Just three specimens added another 18 heterogeneous, technically challenging variants (more than half of the 32 total), and also added 23 additional indels (Tables S1, S6). Importantly, most of these variants were unique, in contrast to variants in the reference and GIAB samples that were often (21% and 74%, respectively) repeated in multiple specimens (repeated variants may be more useful in demonstrating reproducibility compared to sensitivity). Multiple gene panels using different hybridization assays, but an otherwise common workflow, were developed, validated and used in our prevalence study, below.

### Prevalence of technically challenging variants

In our cohort of 471,591 patients meeting study criteria, 102,085 (21.6%) carried one or more clinically reported pathogenic or likely pathogenic (P/LP) variants in 1,217 distinct genes. This positive rate was expected given the mix of tests and clinical indications. A total of 127,710 P/LP variants were reported, of which indels comprised 31.4%, CNVs 9.7%, and SNVs 58.9%. These variants were confirmed as needed^24^ and were thus all confidently considered true positives.

Of these P/LP findings, 13.8% (17,561; 95% CI 13.6 - 13.9%) met one or more of our criteria for being technically challenging (Figure 1). These variants were uncovered in 16,618 patients (i.e., some patients carried more than one) and in 46% of genes (556). As expected, a small number of relatively common variants made up a disproportionate fraction of positive findings: 11 specific sites accounted for 22.2% (28,351) of P/LP variants, and 34% of these (9,683) were considered challenging. At the same time, 18,856 P/LP variants were observed in only a single individual (i.e., these were rare in our cohort), and 9.2% (2,434) of these were considered challenging.

Technically challenging variants were prevalent among all clinical areas studied (Figure 3), particularly in carrier screening, neurology, pediatrics, and hereditary cancer testing, comprising between 10.3% and 20.4% of all P/LP findings in these patients. Prevalence was lower, yet still clinically significant (2.1% to 4.3%), in cardiology, metabolic disorders, preventative testing, immunology and other indications. A list of the genes and findings by type is provided in Table S7.

**Figure 3.**
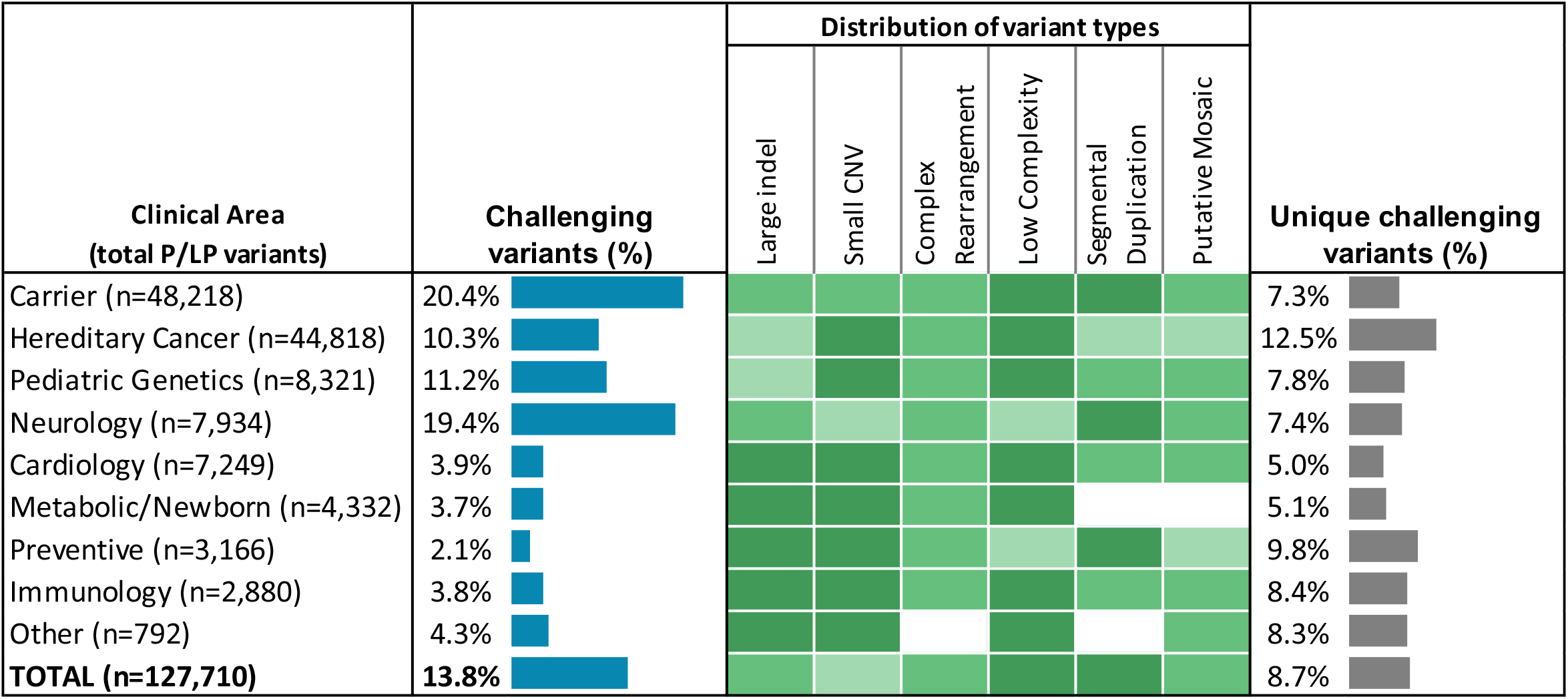
Prevalence of technically challenging variants. For each clinical area, we evaluated the population of pathogenic or likely pathogenic (P/LP) variants that met one or more of our definitions of technically challenging (Figure 1). Blue bars indicate the prevalence of challenging variants among all reported P/LP findings. The heatmap (green cells) indicates the relative contribution of each variant class to this result. Gray bars indicate the fraction of unique variants that were technically challenging (i.e., when the same variant appeared in more than one patient, it was counted only once in this analysis but was counted multiple times in the prevalence analysis [blue bars]). The differences between these two fractions result from a small number of relatively common P/LP variants that are (e.g., in carrier or neurology testing) or are not (e.g., preventative testing) technically challenging. A total of 102,085 patients with P/LP variants in 1,217 genes are represented in this data set. Challenging variants of most types were observed across clinical areas. **Abbreviations**: CNV, copy number variant; Indel, insertion or deletion.

No single attribute defined all or even most of the technically challenging variants we observed. Rather, a broad spectrum was present. Of the challenging P/LP variants, 42.3% (7,423) were located in low-complexity regions (e.g., homopolymers, short tandem repeats) and 35.0% (6,153) were in segmental duplications (segdups). In addition, 11.4% (1,995) were small CNVs, 6.5% (1,135) were large indels, and 6.3% (1,127) were complex rearrangements. Finally, 0.6% of variants (740) were flagged as potentially mosaic based on having an abnormally low NGS allele fraction. (Note that not all of these variants were, in fact, mosaic, and some may be a result of clonal hematopoiesis, but all may warrant investigation). Some variants (118) fell into more than one category (e.g., large indel within a segdup).

A sizable fraction of the low-complexity variants (5,254, 70.8%) were alterations at the *CFTR* intronic poly-T/poly-TG site, which, depending on diplotype, confer modest to moderate risk for pancreatitis, respiratory disease, and male infertility.^31^ Excluding *CFTR*, 2,169 other low-complexity variants were uncovered in 233 different genes, comprising 1.7% of all P/LP findings. Some were particularly challenging for conventional NGS. For example, 91 confirmed findings of *MSH2* variant NM_000251.3:c.942+3A>T were observed, which is not a homopolymer length change, but rather an SNV at the end of a 25 bp homopolymer. This single, high-penetrance pathogenic variant made up 11.0% of all P/LP findings in *MSH2*, a gene conferring cancer risk (Lynch syndrome) as well as response to certain immuno-oncology (IO) drugs.^32^ An additional 185 premutation and full mutation alleles in *FMR1*, underlying fragile X syndrome, were not included in the counts above, owing to methodological differences.

The most common (5,457) findings in segmentally duplicated genes were observed in *SMN1/2, GBA*, and *HBA1/2*. All were tested in carrier screening, with *SMN1/2* also included in neurology tests. Other segdups, including *NEB* (exons 83–103), *PMS2* (exons 12–15), *PRSS1*, and *SDHA*, accounted for 358 findings within the hereditary cancer, neurology, and pediatric indications. For instance, *PMS2* (like *MSH2*) is involved in Lynch syndrome and IO response, and 20.9% of all 1,194 P/LP findings were located in the pseudogene-associated exons. In *NEB*, underlying nemaline myopathy, 7.7% of P/LP variants were in the triplicated exons.

Large indels, small CNVs, and complex rearrangements collectively represented 3,366 P/LP findings, 6.4% of all non-SNVs, affecting 38% of genes and every clinical area (Figure 4a). More than half (1,836) of these were deletions between 50 bp and one exon in size. Whether such events were considered CNVs or indels was, in practice, defined more by methodology than biology. Mobile elements, sometimes called “jumping genes”, accounted for 128 findings, 58 of which were observed in only a single individual.

**Figure 4.**
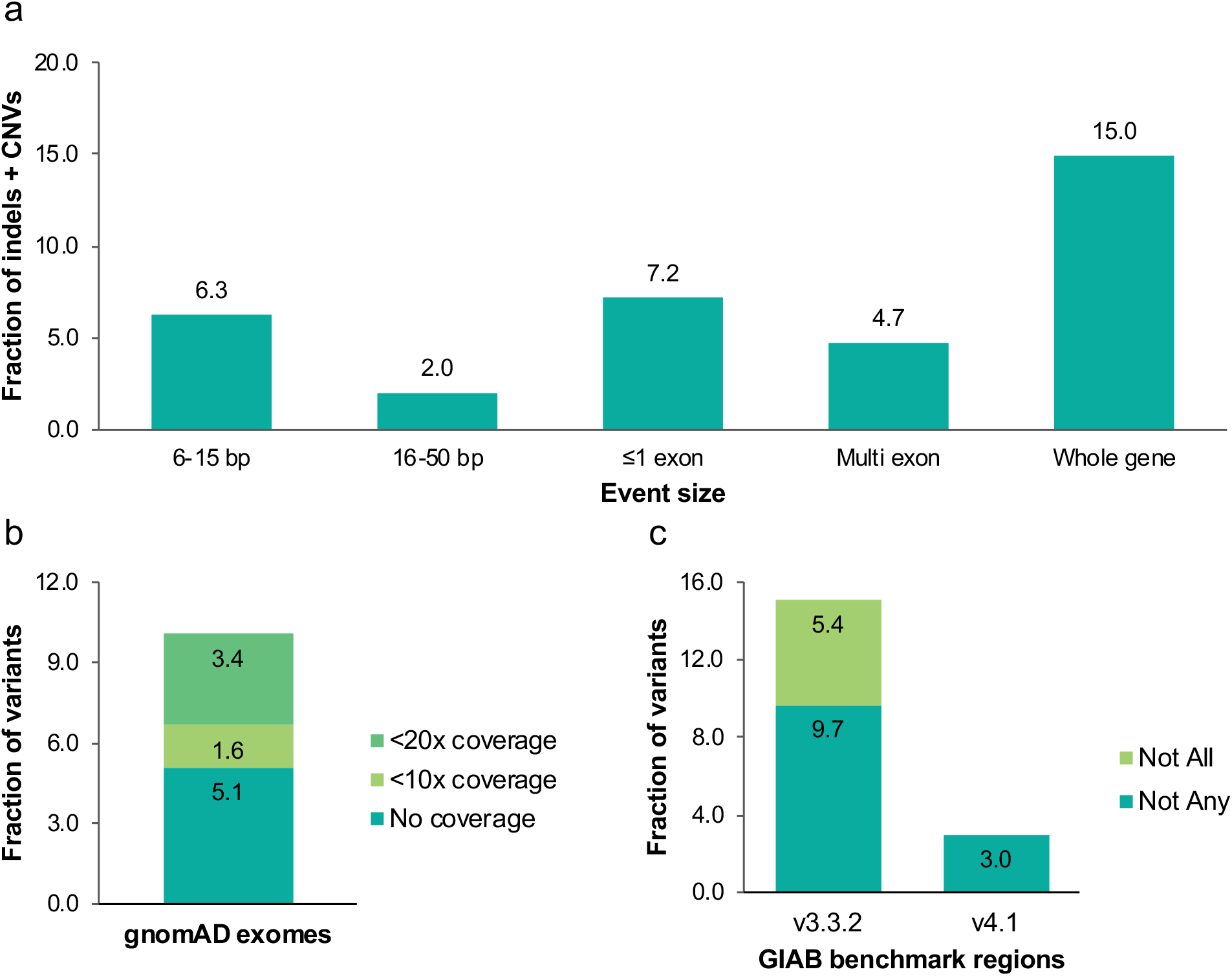
Breakdown of clinical variants. (a) Size distribution of P/LP indels and CNVs, whether technically challenging or not. 64% of these variants were 1-5 bp in size (not shown). SNVs, *FMR1* trinucleotide repeat expansions and variants in the *CFTR* poly-T/TG site are not included. (b) NGS coverage of P/LP clinical variant locations in the gnomAD database of 125,748 whole exome sequences (version 2.2.1). The gnomAD whole genome sequences were not used in this analysis. The average gnomAD exome coverage was 76x at these clinical variant sites (much lower than the 660x average for our clinical testing). The observed rate of a clinical variant location having less than the indicated degree of coverage in the gnomAD exomes was calculated at specific thresholds shown. 5.1% have no coverage (0x), 6.7% less than 10x coverage (including 0x), and 10.1% less than 20x. CNVs were not included in this analysis. (c) Comparison of P/LP clinical variant sites with the Genome in a Bottle (GIAB) benchmark regions using the version 3.3.2 and 4.1 GIAB data sets. Many (9.7%) of these variants were outside of the benchmark regions in all seven GIAB samples (“Not Any” category) and 15.1% of these variants were outside of these regions in at least one of the seven samples (“Not all”). However, the newer version 4.1 GIAB data, available only for one of the GIAB samples at this time, substantially improves this situation. CNVs were not included in this analysis. **Abbreviations:** CNVs, copy number variants; Indels, insertions or deletions; NGS, next-generation sequencing; P/LP, pathogenic or likely pathogenic; SNVs, single nucleotide variants.

### Comparisons with public datasets

As expected, most of the P/LP variants we observed were rare, and thus absent from population databases including gnomAD^33^ (data not shown), although some of these absences are explained by methodological differences in variant detection between gnomAD and our data. Nevertheless, we examined the gnomAD version 2.1.1 exome sequences as a representative, if heterogeneous and research-oriented, view of the coverage that exome capture may achieve at the locations of P/LP variants in our study. Although the average coverage among the 125,748 gnomAD exome sequences at these sites is 76x, our P/LP variants had a 5.1% chance of having no coverage in a gnomAD exome and a 10.1% chance of having <20x coverage (Figure 4b). Even if doubled to ∼150x on average, more typical of clinical exome sequencing, this coverage would likely remain inadequate to detect many of our challenging P/LP variants.

We similarly compared our variant sites with the GIAB benchmark regions for all seven GIAB samples, and found that 15.1% of variants were outside of these regions in at least one of the seven, and 9.7% were outside in all seven using version 3.3.2 GIAB data (Figure 4c). A new release of version 4.1 GIAB data was available for one sample (HG002) in which only 3.0% of our P/LP variants were outside of the benchmark regions, a remarkable improvement resulting from the GIAB consortium’s recent use of both long- and short-read sequencing with improved bioinformatics.^34^

## DISCUSSION

The comprehensive assessment of pathogenic variants is crucial to diagnosis of hereditary disorders and clinical decision making. Positive findings often suggest specific treatment or management actions, and equally the lack of findings can suggest a different course. Thus, both high sensitivity and its corollary, high negative predictive value (NPV), are valued in genetic testing. For example, molecular testing for severe hemophilia A had limited utility until the discovery of *F8* inversions.^35^ Subsequently, accurate tests became available not only for affected patients, but also family counseling, carrier screening, and prenatal diagnosis.^36^

To design and validate (as a lab), or select (as a clinician), the most appropriate tests for any gene or condition, understanding the spectrum of pathogenic variation is important. Our study found that pathogenic variants of technically challenging types for NGS are prevalent across many genes in a large population of patients with diverse clinical indications. These challenging types are heterogeneous and include large indels, small CNVs, mobile element insertions, complex rearrangements, as well as variants within segmental duplications or low-complexity regions. No current NGS platform or current variant calling algorithm can capture all of these variant types. Rather, we found that a battery of algorithms was required and that supplementing short-read NGS with long-read sequencing allowed the resolution of otherwise ambiguous variant calls. Furthermore, we found that NGS methods achieving high sensitivity for some of these variant types also have poor specificity, making confirmatory assays mandatory in some cases, a topic we have detailed separately.^24^ Of course, “challenging” is a relative term, and the specific definition we used (Figure 1) may be too conservative for some laboratory methods, and too permissive for others (Supplement).

The prevalence rates we observed for challenging variants are undoubtedly underestimates, as we know that the NGS workflow used in our prevalence study cannot achieve perfect sensitivity for all variants of all types, despite the validation study results described above. Nor have all of the challenging genes or regions relevant to the clinical areas we studied been implemented into this workflow yet, some of which will require additional methodologies (as did *FMR1)*. Furthermore, there are many clinically relevant genes other than the 1,217 described herein, and non-coding regions of clinical impact continue to be uncovered. Further study of pathogenic variation is clearly needed.

Our interlaboratory study highlighted the fact that the sensitivity of different NGS workflows can vary, particularly based on the bioinformatics pipeline used. While off-the-shelf bioinformatics solutions fared least well in this comparison (Table 1), many of the NGS workflows in this study have improved since, based, in part, on observations from this study. While our studies focused on panel testing, many of the issues we identified would equally apply to whole exome or whole genome sequencing. Unfortunately, we observed that our study’s pathogenic variants may receive low (or no) coverage in some exome sequencing workflows, and coverage variability in exome sequencing may also affect sensitivity (particularly for small CNVs). Carefully evaluating the coverage and limitations of any NGS methodology for each specific application remains vital.

A recent systematic review of validation studies (including both germline and somatic tests) by Roy et al.^3^ found “a clear absence of uniformity” in study design, where (among other issues) highly variable numbers of samples and positive control variants were used to make sensitivity claims. Moreover, some of these studies combined large numbers of SNVs with very few indels, presenting a single measurement of sensitivity with tight statistical bounds. Our studies show that this is often inappropriate, given that indel sensitivity can vary greatly among workflows for all but the smallest and simplest indels, and that indels and/or CNVs can be a substantial fraction of pathogenic variants in many genes. As an example, we have data from one group (not included in Table 1) who performed whole genome sequencing of our synthetic controls and then included benign intronic SNVs in their analysis to argue for >99% overall sensitivity, despite missing many of the sample’s pathogenic variants (including most of the technically challenging ones). As recommended by guidelines,^3^ our results demonstrate that reporting sensitivity and statistical confidence by variant type (SNV, indel, CNV) is critical. Certain guidelines go further, specifying that variants should be broken down by size range,^28^ which our results also support. Highlighting the number (or lack) of low-complexity and segdup-associated variants in a validation study is also critical when such regions are targeted by a test. If a single analytic sensitivity measurement is required, then we believe that it would best be computed on a prevalence-weighted basis, using the known distribution of pathogenic variant types, not based on the distribution of variants that happen to be available for validation. Otherwise, sensitivity estimates may be misleading.

Thoroughly validating NGS workflows can be difficult owing to the paucity of readily obtained positive controls containing nontrivial variants in coding regions of clinically relevant genes. This partially explains the overreliance on SNVs for test validation that Roy et al. observed. In the sensitivity study described herein, reference samples from public biobanks complemented the GIAB resources, although many such samples were required. Clinical specimens with independent data were also informative, although these are not replenishable – new specimens would need to be obtained to re-validate processes. We thus developed a set of replenishable synthetic controls containing diverse challenging variants. These accurately mimicked endogenous variants, showed high utility in our interlaboratory study, and greatly improved the breadth of our sensitivity study. Complementary approaches, such as *in silico* validation^37,38^ are promising for the future, although ongoing development is required to ensure that these approaches capture all of the complications in NGS data (e.g., Figure S1) particularly for challenging variant types. New validation standards and regulatory science advances will be needed to continue the rapid evolution of clinical genetic testing.^39^

In summary, our results demonstrate that clinically significant but technically challenging variants are prevalent in genes associated with a wide-range of clinical indications. As is the case with most genetic diseases, these variants are diverse and often individually rare, but collectively common. Limitations in their detection could lead to appreciable clinical false negative rates. Testing for these variant types is, however, not uniformly implemented, and some validation studies emphasize simpler (and less clinically relevant) variant types. The resources and approaches described herein may help laboratories and clinicians optimize genetic testing to further improve patient care.

## Data Availability

All variants in this study have been submitted to ClinVar.

